# Inferring Skin-Brain-Skin Connections from Infodemiology Data using Dynamic Bayesian Networks

**DOI:** 10.1101/2023.05.15.23290003

**Authors:** Marco Scutari, Delphine Kerob, Samir Salah

## Abstract

**Background:** The relationship between skin diseases and mental illnesses has been extensively studied using cross-sectional epidemiological data. Typically, such data can only measure association (rather than causation) and include only a subset of the diseases we may be interested in.

**Objective:** In this paper, we complement the evidence from such analyses by learning an overarching causal network model over twelve health conditions from the Google Search Trends Symptoms public data set.

**Methods:** We learned the causal network model using a dynamic Bayesian network, which can represent both cyclic and acyclic causal relationships, is easy to interpret and accounts for the spatio-temporal trends in the data in a probabilistically rigorous way.

**Results:** The causal network confirms a large number of cyclic relationships between the selected health conditions and the interplay between skin and mental diseases. For acne, we observe a cyclic relationship with anxiety and attention deficit hyperactivity disorder (ADHD) and an indirect relationship with depression through sleep disorders. For dermatitis, we observe directed links to anxiety, depression and sleep disorders and a cyclic relationship with ADHD. We also observe a link between dermatitis and ADHD and a cyclic relationship between acne and ADHD. Furthermore, the network includes several direct connections between sleep disorders and other health conditions, highlighting the impact of the former on the overall health and well-being of the patient. The average *R*^2^ for a condition given the values of all conditions in the previous week is 0.67: in particular, 0.42 for acne, 0.85 for asthma, 0.58 for ADHD, 0.87 for burn, 0.76 for erectile dysfunction, 0.88 for scars, 0.57 for alcohol disorders, 0.57 for anxiety, 0.53 for depression, 0.74 for dermatitis, 0.60 for sleep disorders and 0.66 for obesity.

**Conclusions:** Mapping disease interplay, indirect relationships, and the key role of mediators, such as sleep disorders, will allow healthcare professionals to address disease management holistically and more effectively. Even if we consider all skin and mental diseases jointly, each disease subnetwork is unique, allowing for more targeted interventions.

## Introduction

Skin diseases and mental illnesses have been extensively studied. However, they are commonly investigated in isolation: the interplay between different skin diseases, between mental illnesses, and between skin diseases and mental illnesses are ignored, limiting our understanding of their aetiology. Skin and brain may interact in four ways: skin-to-skin, brain-to-brain, skin-to-brain and brain-to-skin. Skin-to-skin interactions, or skin disease associations, may arise because of the general altered skin barrier function shared by all inflammatory skin diseases^1^; because of the use of topical drugs that induce an alteration of the skin barrier function or other skin reactions^2,3^; and because of common aggravating mental and environmental risk factors^4,5^.

As for brain-to-brain interactions, there is a growing consensus among psychiatrists that the boundaries between mental disorders, which often overlap in signs and symptoms, are not clear: a recent study shows that mental diseases share a large number of genetic variants^6^. Anxiety and depression, for example, have a genetic correlation of 0.79, while ADHD has a correlation of 0.39 with anxiety and 0.52 with depression. Clinical practice should take into account these overlapping genetic contributions to reduce diagnostic errors and treatment effect heterogeneity on psychiatric disorders.

Skin-to-brain interactions have largely been investigated through the evaluation of the impact of skin diseases on mental health, mainly anxiety, depression and attention deficit hyperactivity disorder^7–13^. This relationship has also been studied indirectly through the impact of skin diseases on symptoms directly related to mental illnesses such as depressive symptoms, social isolation and loneliness^14–16^. A second important pathway is through mediators like the quality of sleep in connection with pruritus^17–20^. Over-representation of sleep disorders has been observed in patients with psoriasis, atopic dermatitis, hidradenitis suppurativa and vitiligo^21–24^. More recent works study the reverse effect of sleep deprivation on skin disease through the bi-directional relationship between sleep and the immune system^25^. This link is thought to contribute to the chronic inflammation observed in many skin diseases. Therefore, dermatologists should emphasise sleep hygiene in their practice.

Dermatologists studying brain-to-skin interactions have been arguing that stress, anxiety and depression can aggravate or precipitate the onset of most inflammatory skin diseases. The COVID-19 pandemic provided strong evidence supporting this hypothesis, with dermatologists reporting an increase in the incidence of flares during this period^26^. Unfortunately, this effect is difficult to quantify because patient access to medical treatment was restricted during lockdown periods when stress, anxiety and depression were most likely to develop. The hypothalamic pituitary adrenal (HPA) axis is responsible for responding to psychological stress, which produces both pro- and anti-inflammatory effects on the skin^4^ in turn. Initially, the release of pro-inflammatory cytokines by the corticotropin-releasing hormone (CRH) starts a quick inflammatory process. However, CRH also triggers a slower anti-inflammatory process that leads to the release of glucocorticoids (cortisone, cortisol). Studies have shown that pro-inflammatory cytokines induce mast cell activation, promoting immune dysregulation and neurogenic inflammation^27,28^. They are known to play a role in allergic reactions as well^29^. Inflammatory skin diseases share a common link with the quality of the skin barrier function: the enzyme 11*β* -hydroxysteroid dehydrogenase type I, which is responsible for the transformation of cortisone (inactive form) into cortisol (active form), is a marker of barrier function impairment^30^. Anti-psychological stress interventions like SSRI reduce both enzyme concentration and improved barrier function.

Psychological stress is not only an aggravating factor for skin diseases but may also trigger diseases like vitiligo^31^, psoriasis^32^, seborrheic dermatitis^33^, trichotillomania, excoriation disorder and delusions of parasitosis^34^ in individuals with genetic susceptibility.

Complementing this epidemiological evidence with a model of the interplay between these four classes of interactions is crucial to improving the diagnosis and treatment of skin diseases and mental illnesses in medical practices. Modelling this interplay using epidemiological data is very difficult because of the lack of comorbidity studies with a longitudinal design. In this paper, we consider the search trends infodemiologic data available in the Google COVID-19 Public Data, an analysis-ready large longitudinal data set^35^. It is not the first time that this data set has been used to complement epidemiological insights for achieving a larger sample size or for disease early detection^36^. The intuition behind its use is that many patients perform online searches about their putative conditions before visiting a physician^36,37^. We can then assume a non-negligible association between the frequency of online searches for specific diseases and the actual incidence of those diseases in physicians’ diagnoses. Restricting ourselves to searches performed on Google is not a significant limitation because of the prevalence of its use: 8.5 billion queries are processed by Google every single day^38^. At the time of this writing, a search of PubMed titles and abstracts with the keywords “search trend” and “Google search” yields 1489 results in 2020, 59% more than in 2019. This volume has been maintained in 2021 and 2022. The COVID-19 pandemic has highlighted how such data can be used to track pandemics and, more interestingly, to study longitudinal patterns of disease progression in several healthcare domains^39,40^. However, poor documentation practices and high heterogeneity in methods have led to conflicting findings^41^. Previous infodemiologic studies based on Google search trends relied on keyword matching, which is likely to miss many classes of queries, and many of them did not incorporate query translation, which may result in significant loss of information in countries like the US where multiple languages are in common use^42^. To overcome these two issues, Google leveraged the latest advancements in natural language processing (NLP) for query classification and translation, including state-of-the-art transformers deep neural networks^43^, to create the COVID-19 Public Data set. Training on large corpora makes them more accurate in classifying diseases even with little to no fine-tuning (respectively, zero-shot classification without additional supervised learning^44^ and with few-shot classification with a small number of examples^45^).

We will use the COVID-19 Search Trends Symptoms data set, which is part of Google’s COVID-19 public data, to model the interplay between skin diseases and mental illnesses with a causal model built on dynamic Bayesian networks (dynamic BNs)^46^. This class of graphical models has a unique set of advantages: they provide a graphical representation of the interactions between diseases; they can be learned automatically from data, from expert knowledge, or a combination of both; and they can be used as diagnostic or prognostic support systems because they can easily evaluate any scenario of interest. Most importantly, unlike the more common correlation networks, dynamic BNs represent interactions as directed arcs that can disambiguate causes and effects^47,48^. This ability is crucial in finding appropriate targets for treatment, thus avoiding symptomatic treatment and improving clinical outcomes, and in differentiating unidirectional cause-effect relationships from feedback loops. As a result, dynamic BNs provide a clearer picture of the interplay between the skin and the brain and, at the same time, they are better suited to design treatment regimes.

## Results

The data set used in this work is publicly available at the URL listed in the Data Availability. We consider the web search queries collected weekly from users in the United States between 2020-03-02 and 2022-01-24 (100 weeks). We focus on the following 12 conditions (with abbreviations): obesity (“OBE”), acne (“ACNE”), alcoholism (“ALC”), anxiety (“ANX”), asthma (“ASTH”), attention deficit hyperactivity disorder (“ADHD”), burn (“BURN”), depression (“DEP”), dermatitis (“DER”), erectile dysfunction (“ED”), sleep disorder (“SLD”) and scar (“SCAR”). We detail the mapping between these conditions and the variables in the search trends in Supplementary Table 1. To avoid a very sparse data set, we remove symptoms with more than 30% missing data. The focus is on skin diseases, excluding very general symptoms like “Lesions”, “Skin conditions”, “Infection”, “Inflammation” and “Skin ulcer” or with potential confusion like “Xeroderma”. The second set of symptoms are mental conditions with a focus on anxiety, depression and ADHD, the most studied mental disease in dermatology. We retain asthma for its documented link with atopic dermatitis and anxiety^49,50^. Obesity is a comorbidity that plays an important role in mental disorders like anxiety^51^, so its presence is justified to control confounding in some relationships. Obesity may also trigger some skin diseases like atopic dermatitis and psoriasis^52,53^. Erectile dysfunction is selected because it is considered a probable consequence of mental disorders and sleep deprivation^54,55^. Considering additional health conditions might allow the network to recover more information but is likely to produce dense networks that are difficult to interpret and make computations cumbersome. For each of these conditions, we retrieved the relative frequency in web search queries of the relevant search terms from the Search Trends Symptoms data set in Google’s COVID-19 Public Data. Frequencies are measured in each county of each US state over the period. Therefore, the resulting data set we used for the analysis is a weekly multivariate time series over 12 health conditions for each county and state with the relative frequencies of the web search queries normalised at the county level. Missing data were imputed, and we confirmed the accuracy of the imputation process to be satisfactory. (See the Methods for details). The overall number of observations for each condition given by 2879 counties over 50 states and 100 weeks is 287900.

The dynamic BN model we learned from these data to investigate the skin-brain-skin causal connections among the conditions above is shown in Figure 1. The model should be read as follows: each node corresponds to one of the conditions above, and arcs represent direct probabilistic associations. Bidirectional arcs represent feedback loops, while unidirectional arcs between different nodes represent one-way effects. Finally, arcs from a node to itself represent each node’s autocorrelation through time. Nodes not connected by an arc are indirectly associated if we can find a path that connects them without passing through any node corresponding to conditions we may be controlling for, or conditionally independent otherwise. The longitudinal data we learned the dynamic BN from allow us to give an additional causal interpretation to the results within Granger’s and Pearl’s causality frameworks, as we will discuss in more detail in the Methods.

**Figure 1.**
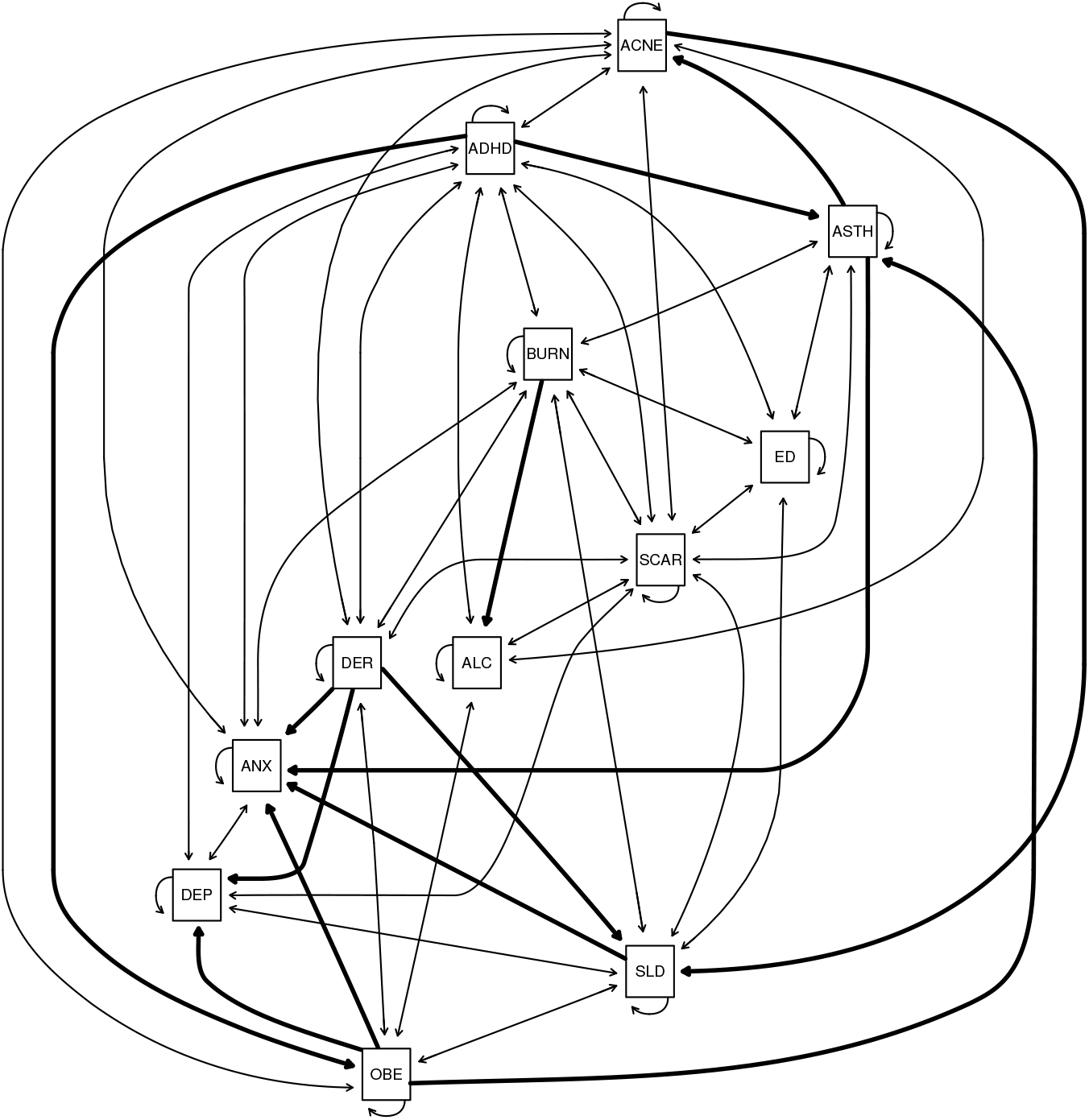
The dynamic Bayesian network linking skin diseases and mental illnesses learned from the Google COVID-19 Search Trends Symptoms public data set. Bidirectional arcs represent cyclic relationships, that is, feedback loops. Unidirectional arcs between different nodes represent one-way causal effects and are drawn with thicker lines for emphasis. Those from a node to itself represent autocorrelation through time.

The interactions between these conditions were learned completely from the data to validate known relationships from the literature and discover new putative ones. To reduce false positives (which implies including spurious arcs in the dynamic BN in this context), we structured the dynamic BN to account for the temporal and spatial dependence structure of the data, and we integrated model averaging via bootstrap resampling in the learning process. Furthermore, we tuned the learning process by penalising the inclusion of arcs in the dynamic BN to find the optimal balance between predictive accuracy and the need for a sparse network. The large sample size of Google’s Search Trends Symptoms data set gives us enough statistical power to detect even marginal effects. However, an overly dense network model would hardly be interpretable from either a qualitative or quantitative point of view: it would be difficult to examine visually, and it would fail to formally identify which pairs of conditions are independent or conditionally independent. Therefore, we chose the final dynamic BN shown in Figure 1 to have about three arcs for each condition. Even so, the average predictive accuracy across all conditions is 99.96% of that of the dynamic BN learned in the same way but with no penalisation. In absolute terms, the *R*^2^ for each condition given the values of all conditions in the previous week are 0.42 for ACNE, 0.85 for ASTH, 0.58 for ADHD, 0.87 for BURN, 0.76 for ED, 0.88 for SCAR, 0.57 for ALC, 0.57 for ANX, 0.53 for DEP, 0.74 for DER, 0.60 for SLD and 0.66 for OBE (average 0.67). This level of predictive accuracy suggests that the dynamic BN in Figure 1 may be a realistic causal model of this set of conditions.

The strengths of the arcs shown in Figure 1 are reported in Supplementary Table 2. They can be interpreted as our confidence that the arcs are statistically significant or as their probability of inclusion in the dynamic BN. As expected, most are close to 1 (where 0 represents a complete lack of confidence, and 1 is the strongest possible confidence). In the case of feedback loops, we observe that both arc directions have approximately the same strength with only two exceptions: acne and dermatitis (1 for ACNE → DER, 0.906 for DER → ACNE), and ADHD and dermatitis (0.688 for ADHD → DER, 0.996 for DER → ADHD). The difference in arc strength was 0.026 or less for all other feedback loops. Note that the model allows us to give a causal interpretation to the arcs within the frameworks of Granger’s and Pearl’s causality, so we can interpret these arc strengths as the probability of causal effect between the variables they connect.

We show in Figure 2 the proportions of variability of ACNE, ANX, DER, DEP, SLD and ADHD explained by the respective parents, normalised by the total variance explained for the condition: these conditions are of particular interest to us. (The raw proportions are shown in Supplementary Figure 1.) The proportion of variability explained by each parent measures the magnitude of its direct causal effect on the trait and complements the corresponding arc strength (which only measures how confident we are that a causal effect exists at all). This information is especially useful in the case of the dynamic BN from Figure 1 because all the arc strengths are equal to or close to 1, regardless of the associated effect sizes, because of the large sample size of Google’s Search Trends symptoms data set.

**Figure 2.**
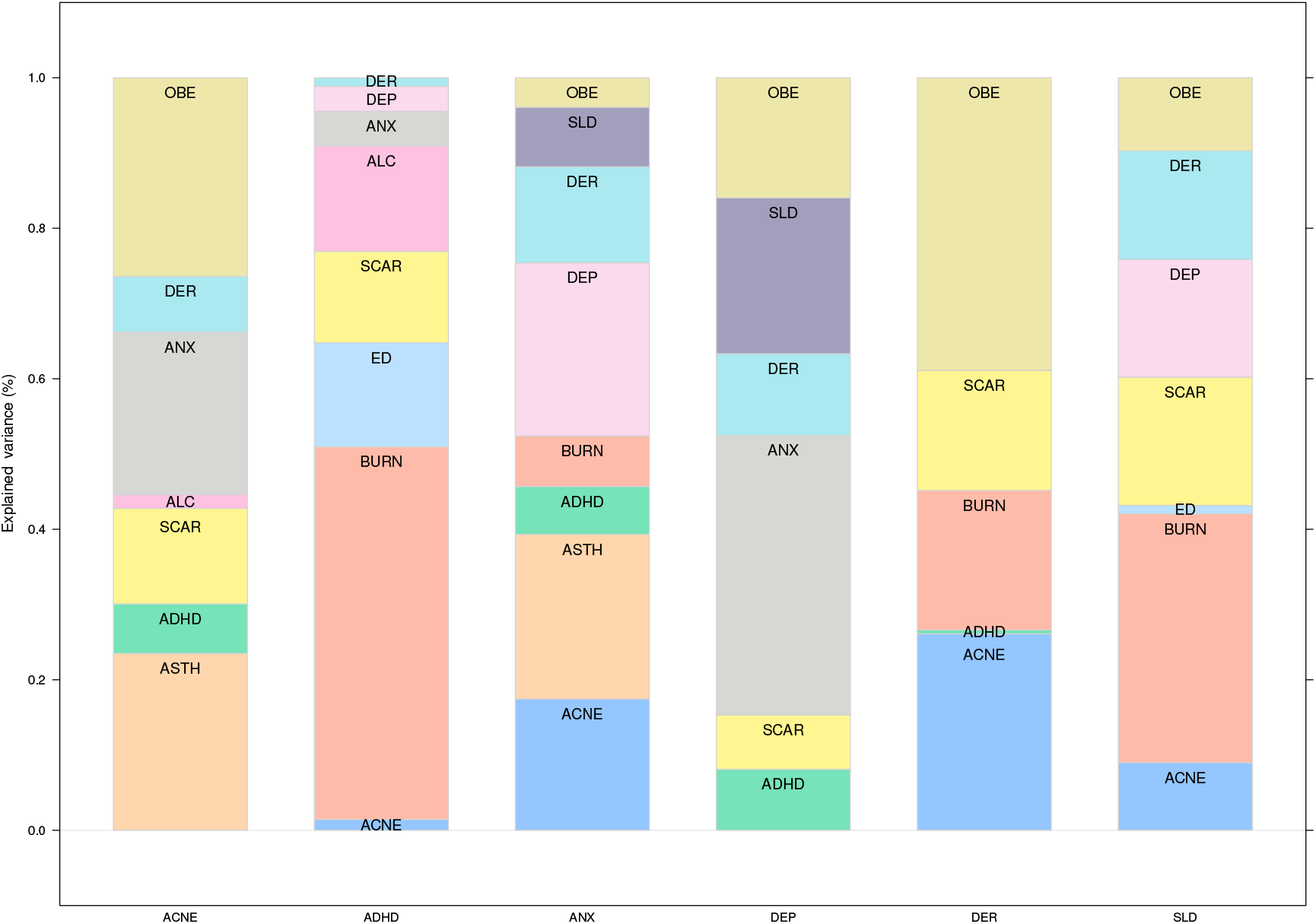
Proportions of the variance of some conditions of interest explained by their parents in the network shown in Figure 1, normalised by the total explained variance. The proportions are computed excluding the contribution of the condition itself from the previous time point in the data.

In particular, we note that ASTH accounts for a nontrivial share (23.5%) of the explained variability of ACNE in the following week, less than OBE (26.4%) but more than ANX (21.6%). However, the association between the ASTH and ACNE is not well established in the literature. We attribute this finding to the fact that ANX is connected to all of ASTH, ACNE and OBE, which may introduce confounding in models that do not consider them simultaneously. For instance, such confounding may arise because of the feedback loop between ACNE and OBE (which are both connected to ANX) unless OBE is controlled for. Our model supports the hypothesis that ASTH and ACNE are only weakly associated (in the same week) after controlling for ANX (in the previous week): if we perform inference on the dynamic BN as described in the Methods, ASTH only accounts for 0.41% of the explained variance of ACNE. Stratifying by ANX (low: bottom quartile of search query frequencies; high: top quartile; average: second and third quartiles) reveals that the share of explained variance is even smaller when ANX is average or low (0.46%, 0.36%, respectively) compared to when ANX is high (0.53%).

Furthermore, the model supports the hypothesis that the feedback loop between ACNE and OBE is driven by ANX: ACNE account for a sizeable share of the explained variability of OBE (62.2%) and vice versa (26.4%). However, after controlling and stratifying for ANX using BN inference, OBE only accounts for a trivial share of the explained variability of ACNE in the same week (1.6% for high ANX, 1.7% for low ANX and 1.3% for average ANX). The same is true for ACNE, which accounts for similar share of the explained variance of OBE.

These results confirm the cyclic relationships between skin diseases and mental illnesses. Figure 2 highlights the importance of each trigger for the six health conditions. Depression is mainly driven by mental disorders and sleep disorders, but dermatitis explains a significant proportion of its search popularity. Anxiety triggers are more diverse: skin conditions like acne and dermatitis play an important role. Triggers of ADHD, learned by the dynamic BN without any reference to clinical experts or the literature, give a new insight into the disease. It is already known that ADHD is associated with a risk increase of burn injuries and scarring (from burns): these data suggest that an increase in burn injuries may lead to ADHD diagnosis.

For acne, we observed a strong, direct cyclic relationship with anxiety and ADHD and an indirect relationship with depression through sleep disorders. For dermatitis, we observed directed links to anxiety, depression and sleep disorders and a cyclic relationship with ADHD. We also observed a link between dermatitis and ADHD, and a cyclic relationship between acne and ADHD. Although not exclusive, the role of mediators like sleep disorders is confirmed in the network with a significant contribution to anxiety and depression. Acne, burns, scars and dermatitis directly impact sleep disorders. The learned BN visualises multiple disease relationships in a single picture.

## Discussion

Our results confirm the interplay between skin diseases and mental illnesses at the infodemiological level. Several skin-to-skin, brain-to-brain, skin-to-brain and brain-to-skin relationships are highlighted in the model. It is interesting to see well-known clinical relationships reproduced in the dynamic BN and put into a larger context with deeper explanations. The data we used for the study uniquely allow for these results by providing almost complete coverage of the US population, with high-quality labelling of web searches, and for an extended period of time.

At the same time, our analysis has two main limitations. Firstly, the correspondence between infodemiological and epidemiological information is a crucial assumption that is difficult to test in practice but that appears to be supported by the other studies using the Search Trends Symptoms data^56–58^. Even so, we cannot draw clinical conclusions about single individuals because the causal relationships we observe at the level of the aggregate data may not necessarily hold for individuals^59^. Secondly, there may be latent confounders that introduce spurious arcs or causal directions in the dynamic BN. The almost complete coverage of the web searches of the US population suggests that neither sampling bias nor systematic patterns of missing values affect our causal analysis. Likewise, latent variables that act as confounding factors at the level of individuals are not necessarily confounding factors at the level of the aggregate data we are analysing. However, there may be conditions that we have not included in the analysis that introduce confounding at the level of the aggregate data. There are also patterns of confounding specific to aggregate data we should be aware of^60^.

A directed link between two conditions in a dynamic BN can represent different types of associations. It may indicate direct causation, where one condition directly influences the other. Alternatively, the link could reflect a temporal dependency without causation, where the two conditions are closely associated in time due to shared genetic factors or similar environmental exposures. Such temporal associations might lead to the sequential manifestation of symptoms, which could be mistakenly perceived as a causal link. Additionally, the observed relationship could be the result of an unobserved confounding factor, introducing bias into the interpretation. It is important to recognise that one condition may not cause the other to occur; rather, the link could suggest that one condition may activate or exacerbate symptoms of an already existing condition.

To illustrate the types of associations, we can consider the arc from ADHD to asthma. This link could imply direct causation if ADHD-related behaviours, such as hyperactivity, or treatments aggravate pre-existing asthma symptoms^61,62^. On the other hand, the link might reflect a temporal association without direct causation, where ADHD and asthma appear together more frequently due to shared genetic markers^63^ or environmental factors^64^ without one condition causing the other, although ADHD symptoms may manifest first. Additionally, an unobserved factor like socio-economic status could confound the relationship, suggesting a link where none exists^65,66^.

Nevertheless, the large number of feedback loops supports the existence of vicious circles in which diseases exacerbate each other until treated appropriately. The dynamic BN cannot elucidate the starting point of these circles but emphasises the need for more holistic disease management for dermatologists and psychiatrists. Dermatologists should take into account the mental health of their patients, and psychiatrists should take into account the skin problems of their patients.

The results also highlight the essential role of key mediators, like sleep disorders, that establish a bridge between the skin and the brain. We should not ignore these disorders if we want to act effectively on skin and mental health. Furthermore, not controlling for comorbidities like obesity may lead to spurious conclusions, hiding the existing relationships.

In addition, the dynamic BN has confirmed the genetic correlation between depression, anxiety, and ADHD^6^, illustrated through bi-directional arcs. The fact that these disorders share numerous signs and symptoms may lead to additional spurious associations, potentially due to diagnostic errors^6^. In the case of acne and dermatitis, these inflammatory diseases are linked by their common feature of altered skin barrier integrity^1^, and they may be triggered by similar environmental conditions^4,5^. Acne treatments are also recognised for their potential to irritate the skin^2,3^. While the direct link between acne and sleep disorders has been less explored than that between dermatitis and sleep disorders, some studies suggest a connection between the severity of acne and sleep disorder^67^. As our data are continuous, we can capture these kinds of associations.

A less evident bi-directional relationship is acne with scars. The acne-to-scars direction is evident and known as severe forms of acne, such as nodules and cysts, can lead to skin damage and subsequent scarring^68^. The scars-to-acne direction is much less evident, but explainable: scar healing can lead to itchiness and may cause irritation and rash^69,70^. Such skin reactions can result in the formation of papules, a situation often seen with hypertrophic scars and keloids.

Even if we consider all skin and mental diseases jointly, each disease subnetwork is unique, allowing for more targeted interventions. The conditional independence property of BNs allows for this kind of focus without loss of information.

In this work, we also wanted to raise awareness of the importance of measuring causality with appropriate study designs and statistical methods leveraging multiple conditions in longitudinal monitoring and allowing feedback loops to reproduce the natural cycle of human health. This may significantly reduce the number of measured associations and highlight a focused set of preferential targets for intervention.

The second important objective of this study was to provide guidelines for better use of search trends data to ensure robust findings. Firstly, query classification using a keywords approach may fail to capture relevant information, leading to low reproducibility across researchers. Using the latest AI breakthroughs in natural language processing for query translation and classification will ensure better reproducibility of studies.

Secondly, choosing models that can capture the main features of search trends data is necessary to avoid several sources of bias. We provided a detailed methodology to deal with missing data, space and time dependencies, the lack of sparsity in the network due to the size of the data set, model interpretability and other considerations.

The marked discrepancies between the conclusions of studies dealing with this type of data can be attributed to how queries were classified and processed. Standardising these two tasks will demonstrate the high potential of these data to complement clinical evidence for a more positive impact on public health.

## Materials and Methods

All analyses were performed using R^71^ and the packages nlme^72^, lme4^73^, imputeTS^74^ and bnlearn^75^.

### The Search Trends Symptoms data

Google released the Search Trends Symptoms data^76^ in September 2020 as part of the COVID-19 Public Data sets. It includes aggregated county- and state-level Google search frequencies for 422 symptoms and conditions that might be related to COVID-19. This data set is unique in that, as mentioned in the Introduction:

- It was aggregated both by day and by week, and normalised by scaling the symptom search count with the total search activity in the county or state. Therefore, it does not require further preprocessing to make it homogeneous across time and space. Furthermore, the data is stationary because of the normalisation.
- Search terms were disambiguated and aggregated across languages and synonyms using Google’s NLP models^43^.
- Missing data in the weekly frequencies were imputed without loss of information from the daily frequencies wherever possible^77^.
- The prevalence of Google’s use in the US^38^ reduces the risk that its web searches are a biased sample that is not representative of those of the US population.

As a result, it has proven an effective tool to construct predictive models for various conditions in infodemiology^56–58^. This is not surprising because 95% of Americans use the Internet^78^, and of those 87.2% use Google: the Search Trends Symptoms data set is almost a population-level longitudinal data set. Therefore, we can consider it unaffected by common sources of sampling bias such as age, race, gender, socio-economic status, or geographical location.

Specifically, we considered the weekly search frequency data collected in the US at the county level from March 2, 2020, to January 24, 2022. This subset of the data spans 2879 counties and 100 time points (weeks) and contains no missing data. Therefore, it takes the form of a multivariate time series over the 12 conditions we are studying (OBE, ACNE, ALC, ANX, ASTH, ADHD, BURN, DEP, DER, ED, SLD and SCAR) for each county. These conditions are completely observed in the chosen time period, which allows us to rule out confounding due to systematic missingness.

### Missing data management

The Search Trends Symptoms data set originally contained missing data, either single data points or whole time series for specific counties and periods. The proportion of missing data in individual conditions ranged from 0% to 16%. We explored different methods to impute them for both individual time series (exponentially weighted moving average, interpolation) and multivariate time series (Kalman filters) using the imputeTS R package. To assess their performance, we used the complete observations and artificially introduced 2%, 5%, 10% and 20% missing values individually and in batches of four to simulate both random missingness and lack of measurements over one month. All combinations of missing data patterns, proportions, and algorithms produced average relative imputation errors between 10% and 14%, suggesting that all imputation methods perform similarly (Supplementary Figure 2 and Supplementary Figure 3). Finally, we chose an exponentially weighted moving average imputation because of its simplicity. Imputing some combinations of conditions and counties was impossible because of the insufficient number of observed values; we dropped them from the data, reducing the overall sample size to 287866.

### Spatio-Temporal dependence structure of the data

The Search Trends Symptoms data set was collected over time and in different US states and counties and, therefore, presents marked spatio-temporal patterns of dependence between observations. We will summarise them here because they are crucial in our modelling choices.

To be consistent with the assumptions of the dynamic Bayesian network we will describe below, we quantify the magnitude of spatial and temporal dependencies with the proportion of the variance of the conditions that they explain as random effects in a mixed-effect model^72^. For spatial dependencies, we further distinguish between the variance explained by the state and by the county. For time, we assume an autocorrelation of order 1 (that is, each condition is correlated with itself at the previous time point). The proportions for each condition and the average over all conditions are shown in Supplementary Figure 4. On average, states explain 12% of the variance of the conditions (min: 7%, max: 16%), counties explain 49% (min: 23%, max: 84%), and counties together with autocorrelation explain 64% (min: 35%, max: 86%).

### Dynamic Bayesian networks

Bayesian networks (BNs)^46^ are a graphical modelling technique that can leverage data combined with an expert’s prior knowledge to learn multivariate pathway models. The graphical part of the BN is a directed acyclic graph (DAG) in which each node corresponds to a variable of interest (here, one of the conditions) and in which arcs (or the lack thereof) elucidate the conditional dependence (independence) relationships between the variables. The implication is that each variable depends in probability on the variables that are its parents in the DAG: as a result, the joint multivariate distribution of the data factorises into a set of univariate distributions associated with the individual variables. This property allows automatic and computationally efficient inference and learning of BNs from data and has made them popular for analysing clinical data^79–81^. In particular, BN inference can automatically validate arbitrary hypotheses: in the simplest instance, a BN is used as a generative model, and hypotheses are validated by stochastic simulation.

To account for the spatio-temporal nature of the data, we use a particular form of BN called a dynamic BN in which each variable appears in the DAG as a separate node at each time point. The key advantage of dynamic BNs is that, unlike classical BNs, they can represent feedback loops by allowing each variable in a pair to depend on the other across time points: for instance, 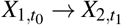 and 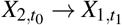 implies a feedback loop between *X*_1_ and *X*_2_ across two consecutive time points *t*_0_ and *t*_1_. Arcs are only allowed to point forward in time from a variable measured at *t*_0_ and one measured at *t*_1_ to ensure that they are semantically meaningful and to be able to relate the dynamic BN to the Granger^47^ and Pearl^48^ causality frameworks. We disallow “instantaneous arcs” between variables measured in the same time point for similar reasons. With these restrictions, we can construct a directed cyclic graph that can represent feedback loops from the DAG by folding the nodes corresponding to the same variable at different time points into a single node. As a result, pairs of arcs like 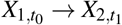 and 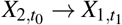 are transformed into cycles of the form *X*_1_ ⇆ *X*_2_; and arcs like 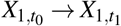 become loops that express autocorrelation. In principle, we could extend this construction to consider arcs from time points further in the past, such as 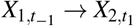. However, the resulting dynamic BN would quickly lose interpretability because the number of nodes (arcs) would increase multiplicatively (exponentially) and because we would lose the ability to display an interpretable cyclic graph.

We should also critically examine the theoretical assumptions underlying causal inference in BNs: the faithfulness condition and the absence of latent confounders. The faithfulness condition requires that the observed probabilistic dependencies are entirely due to the causal structure of the network. The lack of latent confounders, defined as unobserved variables that are parents of at least two observed variables, avoids the risk of edges representing spurious causal effects. Furthermore, the data we learn the BN from should be representative of the population we would like to study and in sufficient quantity to ensure that we have enough statistical power to identify causal effects. They should also be free from sampling bias and systematic patterns of missing values: both act as latent confounders.

As for the distributions of the variables, we assume that 1) the search query frequencies in any given week can depend in probability on those in the previous week but not on older ones, and 2) the data are stationary, so we only need to model the dependence between two generic consecutive times *t*_0_ and *t*_1_. These assumptions allow us to parameterise the dynamic BN similarly to a vector auto-regressive time series^82^: each condition *X*_*i*_ in each county is therefore modelled using a linear regression model of the form

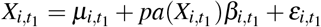

at time *t*_1_ and

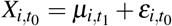

at time *t*_0_. Here *pa* (·) denotes the parents of the variable, *µ*_*i*_ is the intercept, the *β*_*i*_ are the associated regression coefficients, and the *ε*_*i*_ are normally distributed residuals with mean zero and some variance specific to each node. The contribution of each parent to the linear regression can thus be naturally measured by the associated explained variance in the model (ANOVA). Conditions have different scales arising from the different popularity of the corresponding search queries: to make them easier to compare in the Results, we normalise the variance explained by each parent into a proportion (that is, we divide it by the total explained variance of all parents). In doing so, we omit the contribution of the condition itself: in the auto-regressive model we are considering, auto-correlations are strong enough to make the contributions of other conditions appear less significant for purely numerical reasons.

In addition to accounting for the time dimension, the dynamic BN incorporates the spatial structure of the data. Assume that each condition has a different baseline value in each county that does not change over time. Then the local distribution of each condition at *t*_1_ is regressed against itself at *t*_0_ (same county, previous time point). The different baseline for the state then appears on both sides of the equation and can be accounted for in the regression model.

We learned the dynamic BN from the data by choosing the optimal DAG that maximises the penalised log-likelihood

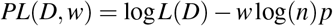

where *p* is the number of parameters of the dynamic BN and *n* is the number of observations in the data, using the hill-climbing greedy search algorithm^83^. We then estimated the parameters of the dynamic BN (the intercepts *µ*_*i*_ and the regression coefficients *β*_*i*_) with the chosen DAG using maximum likelihood. To ensure that the DAG is as sparse as possible without sacrificing predictive accuracy, we chose the penalty coefficient *w* by learning the dynamic BN with *w* = 1, 2, 4, 8, 16, 32, 64, 128 on the first 52 weeks of data and then computing the average proportion of variance explained over all conditions in the remaining weeks of data acting as a validation set. For reference, *w* = 1 gives the Bayesian Information Criterion (BIC)^84^. Larger values of *w* penalise the inclusion of arcs in the DAG by increasingly large amounts, effectively decreasing the value of the penalised likelihood if the associated regression coefficients are small.

The resulting proportions of explained variance are plotted against *w* in Figure 3. We observe no marked decrease in predictive accuracy until *w* = 4, which we choose as the best trade-off with the sparsity of the DAG. For reference, the dynamic BN learned with *w* = 1 has 123 arcs, while that learned with *w* = 4 has 87 arcs with a predictive accuracy that is 99.96% of the former model. We also note that the variance explained by the dynamic BNs in the validation set is not markedly different from that in the training set they were learned from, suggesting no overfitting.

**Figure 3.**
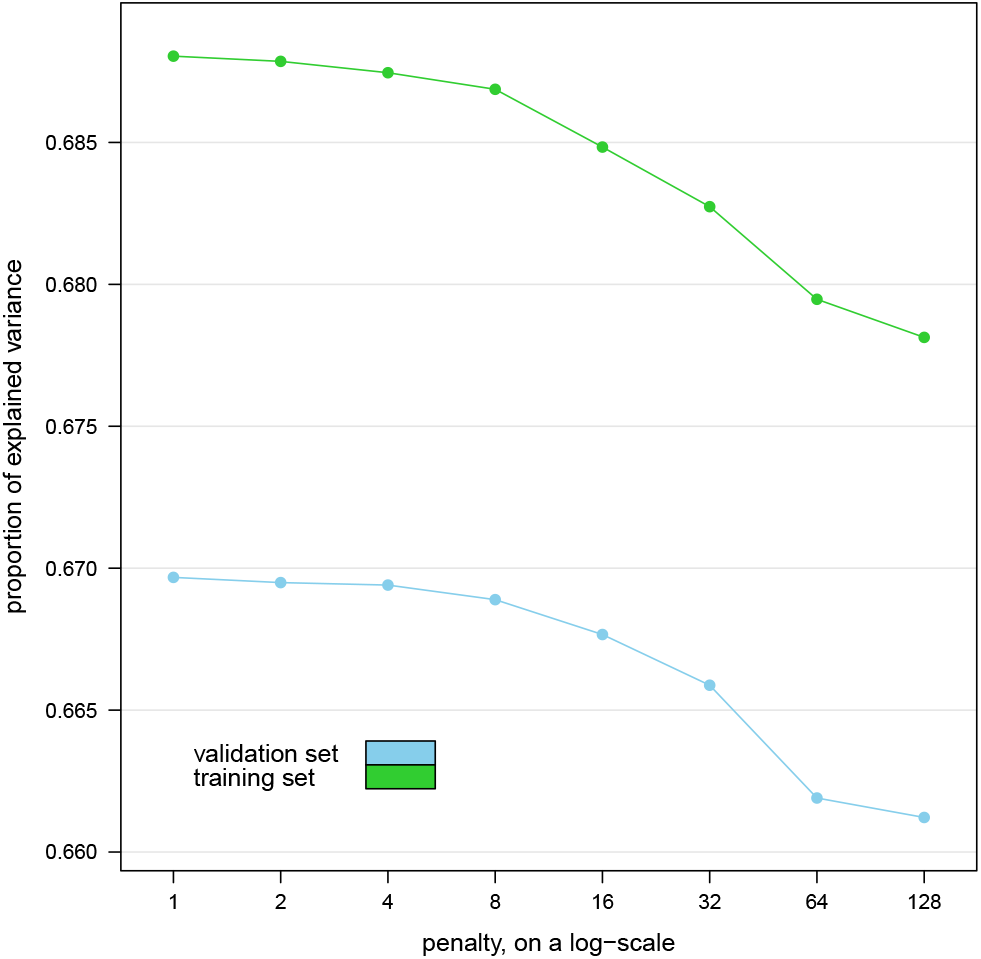
Average proportion of explained variance over all conditions explained by the dynamic Bayesian networks learned from the first year’s worth of data (training set) in the remaining weeks (validation set) for various values of the penalty coefficient *w*.

The large number of observations available on each condition gives the dynamic BN enough statistical power to detect even marginal effects from the data and thus to include them as arcs in the model, resulting in an overly dense DAG. The fact that increasing *w* and dropping many of those arcs has a limited impact on predictive accuracy suggests that the relationships they express may be of limited clinical relevance for diagnostic or prognostic purposes.

Furthermore, we used model averaging to reduce the potential of including spurious arcs in the BN. We implemented it using bootstrap aggregation: we extracted 500 bootstrap samples from the data, learned the DAG of a dynamic BN from each of them, and then constructed a consensus DAG containing only those arcs that appear with frequencies higher than the threshold 0.518 estimated from the data^85^. Each frequency measures the strength of the corresponding arc, that is, of our confidence that the arc is supported by the data: a value of 0 represents a complete lack of confidence, whereas a value of 1 represents perfect confidence. Overall, the individual frequencies are estimated from the joint arc frequencies for all possible combinations of arcs, and they are used to compute the optimal threshold so that we do not have to manually choose its value. If two nodes are connected with a frequency greater than the threshold, the consensus DAG will contain an arc connecting them. The direction of that arc will be the most common direction observed in the DAGs from the bootstrap. Arcs are included in the consensus DAG in decreasing order of strength, and lower-strength arcs are discarded if they introduce cycles. To further prevent patterns in the data from impacting the consensus model, we increased the variability of the bootstrap samples by randomising the order of the variables and by reducing their size to 75% of that of the original data.

Finally, we assessed the impact of the spatio-temporal structure of the data on structure learning to motivate using dynamic BNs. For this purpose, we performed both structure and parameter learning as described above to fit a classical (that is, static) BN in which variables are not replicated across time points. As a means of comparison, we learned a second static BN from the data after removing their spatio-temporal structure with the mixed-effect model we used above to quantify the proportion of variance explained by the counties and the temporal autocorrelation. If we only perform parameter learning, we find that 63% of the regression coefficients are inflated by a factor of at least two compared to those we learn after removing the spatio-temporal structure from the data. The sign is different for 29% of them. If we perform structure learning from both sets of data, we find that 71% of the arcs differ between the two. The reason is that a classical, static BN is a misspecified model, and the dependence relationships between the variables are confounded with space-time dependencies between the data points. If we take the dynamic BN in Figure 1 to be the correct model, the static BN learned from the raw data has 11% correct arcs (*X*_*i*_ → X_j_ in both networks), 50% arcs that should be feedback loops (*X*_*i*_ → *X*_*j*_ in the static network, *X*_*i*_ ⇆ *X*_*j*_ in the dynamic BN), 8% reversed arcs (*X*_*i*_ → *X*_*j*_ in the static network, *X*_*i*_ ← *X*_*j*_ in the dynamic BN) and 30% spurious arcs (*X*_*i*_ → *X*_*j*_ in the static network, no arc between *X*_*i*_ and *X*_*j*_ in the dynamic BN). The proportions for the static BN learned from the data after removing the spatio-temporal structure of the data are similar. This supports our choice to use a dynamic BN: the large number of feedback loops in the causal structure of the data means that any model that cannot express them will report a large number of incorrect causal directions.

## Code availability

The code used for the analysis is publicly available at the URL: https://www.bnlearn.com/research/loreal21

## Data availability

The Google COVID-19 Public Data Set is publicly available at the URL: https://console.cloud.google.com/marketplace/product/bigquery-public-datasets/covid19-public-data-program

In particular, the Search Trends Symptoms data set is available at the URL: https://console.cloud.google.com/marketplace/product/bigquery-public-datasets/covid19-search-trends and at the URL: https://github.com/google-research/open-covid-19-data/

## Acknowledgements

We thank Dr. Katrina Abuabara for her comments and suggestions on an early draft of this paper.

## Author contributions statement

M.S. analysed the data and wrote the software for the analysis. D.K. conceived the study and interpreted the clinical results. S.S. conceived the study, collected the data and interpreted the clinical results. All authors wrote and reviewed the manuscript.

## Competing interests

The authors declare no competing financial or non-financial interests.

## Supplementary information

**Supplementary Table 1** Mapping between the variables in the Google COVID-19 Public Data Set and the conditions discussed in this paper. When multiple variables map to the same condition, the search query frequencies from those variables were aggregated to give a single overall frequency for the condition.

**Supplementary Table 2** Arc strengths for the dynamic Bayesian network model shown in Figure 1. “From” denotes the node at the tail of the arc, “To” denotes the node at the head of the arc, and “Arc strength” is the frequency of the arcs in the bootstrapped models.

**Supplementary Figure 1** Proportions of the variance of ACNE, ADHD, ANX, DEP, DER, and SLD explained by their parents in the network shown in Figure 1, unnormalised. This figure complements Figure 2 in which the proportions are normalised by the total explained variance for the condition.

**Supplementary Figure 2** Average relative error (in absolute value) for the missing data imputation algorithms with individual missing values amounting to 2%, 5%, 10% and 20% of the total.

**Supplementary Figure 3** Average relative error (in absolute value) for the missing data imputation algorithms with values missing in 1-month batches (4 consecutive weeks) amounting to 2%, 5%, 10% and 20% of the total.

**Supplementary Figure 4** Proportion of the variance of each condition explained by the US states, by the counties and by the counties together with the temporal autocorrelation. The average for each of them over all conditions is reported at the bottom.

## Notes

### Competing Interest Statement

The authors have declared no competing interest.

### Funding Statement

This study did not receive any funding

### Summary of Updates

Revised the manuscript upon resubmission, clarifying the assumptions between the causal network model.

